# Large-scale cross-sectional seroepidemiologic study of COVID-19 in Japan: Acquisition of herd immunity and the vaccines’ efficacy

**DOI:** 10.1101/2022.01.13.22269203

**Authors:** Zhenxiao Ren, Mitsuhiro Nishimura, Lidya Handayani Tjan, Koichi Furukawa, Yukiya Kurahashi, Silvia Sutandhio, Kaito Aoki, Natsumi Hasegawa, Jun Arii, Kenichi Uto, Keiji Matsui, Itsuko Sato, Jun Saegusa, Nonoka Godai, Kohei Takeshita, Masaki Yamamoto, Tatsuya Nagashima, Yasuko Mori

**Author notes:** **Corresponding author:** Prof. Yasuko Mori, Division of Clinical Virology, Center for Infectious Diseases, Kobe University Graduate School of Medicine, 7-5-1 Kusunoki-cho, Chuo-ku, Kobe, Hyogo 650-0017, Japan. **Alternate author:** Dr. Mitsuhiro Nishimura, Division of Clinical Virology, Center for Infectious Diseases, Kobe University Graduate School of Medicine, 7-5-1 Kusunoki-cho, Chuo-ku, Kobe, Hyogo 650-0017, Japan.

## Abstract

**Background:** The COVID-19 pandemic situation has been changing drastically worldwide due to the continuous appearance of SARS-CoV-2 variants and the roll-out of mass vaccination. Periodic cross-sectional studies during the surge of COVID-19 cases is essential to elucidate the pandemic situation.

**Methods:** Sera of 1,000 individuals who underwent a health check-up in Hyogo Prefecture Health Promotion Association clinics in Japan were collected in August and December 2021. Antibodies against SARS-CoV-2 N and S antigens were detected in the sera by an electrochemiluminescence immunoassay (ECLIA) and an enzyme-linked immunosorbent assay (ELISA), respectively. The sera’s neutralization activities for the conventional SARS-CoV-2 (D614G), Delta, and Omicron variants were measured.

**Results:** The seropositive rates for the antibody against N antigen were 2.1% and 3.9% in August and December 2021 respectively, demonstrating a Delta variant endemic during that time; the actual infection rate was approximately twofold higher than the rate estimated based on the polymerase chain reaction (PCR)-based diagnosis. The anti-S seropositive rate was 38.7% in August and it reached 90.8% in December, in concordance with the vaccination rate in Japan. In the December cohort, 78.7% of the sera showed neutralizing activity against the Delta variant, whereas that against the Omicron was much lower at 36.6%.

**Conclusions:** These analyses revealed that herd immunity against SARS-CoV-2 including the Delta variant was established in December 2021, leading to convergence of the variants. The low neutralizing activity against the Omicron variant suggests the need for the further promotion of the prompt three-dose vaccination to overcome this variant’s imminent 6th wave in Japan.

**Summary:** Seroepidemiologic study of COVID-19 on December 2021 in Japan showed neutralizing antibodies for Delta were 78.7%, indicating the acquisition of herd immunity by mass vaccination leading to convergence while those for Omicron were only 36.6%, indicating need of booster vaccination.

## INTRODUCTION

Several turning points have occurred since the COVID-19 pandemic’s emergence in December 2019, and the pandemic has undergone drastic changes with its progress. One of the most important factors is the appearance of the continuous SARS-CoV-2 variants replacing the original variant. In Japan, the Alpha variant (B.1.1.7) replaced the existing strain by around April 2021; the Delta variant (B.1.617.2) then began spreading rapidly throughout the country from July to August 2021(it was called the 5th wave in Japan, Suppl. Fig. S1) [1-3]. The Omicron variant (B.1.1.529) was first reported in South Africa in November 2021 [4] and has spread worldwide [5] including UK and U.S. (new Omicron infections: >200,000/day in the UK [6] and >1,000,000 Omicron cases per day in the U.S. [7]), and it was expected to invade Japan at the end of 2021.

The other critical event in the pandemic has been the launch of COVID-19 vaccines. The accelerated development of several vaccine platforms was based on the components of the original SARS-CoV-2 strain as the template, and these vaccines have been demonstrated to be effective for reducing the COVID-19 outbreak [8, 9]. The BNT162b2 mRNA vaccine (Comirnaty^®^, BioNTech-Pfizer, Mainz, Germany/New York, USA) [10] was approved first in Japan and administered to healthcare workers starting in February 2021, then to individuals aged ≥65 old beginning in April 2021, expanding to other populations from May 2021. The mRNA vaccines developed by Moderna [11] were also approved in Japan and have been widely used since May 2021. The number of vaccinated individuals in Japan is increasing rapidly, as monitored by the government system [12]; as of December 2021, approx. 74% of the population has received two doses of a vaccine. A three-dose vaccination policy for medical staff was initiated in December 2021 [13].

Periodic seroepidemiologic surveillance is useful to help determine the precise COVID-19 situation [14-18]. We have periodically conducted seroepidemiologic surveillance in one of Japan’s 47 prefectures; i.e., Hyogo prefecture (population 5.4 million) located in the southern-central region of Japan [19, 20]. Our previous seroepidemiologic surveillance conducted in October 2020 revealed that 0.15% of 10,377 sera had neutralizing activity against SARS-CoV-2 infection[20].

In the present study, we conducted seroepidemiologic surveys of COVID-19 in Hyogo prefecture in August and December 2021, which respectively correspond to the beginning and the end of the pandemic’s 5th wave in Japan. We observed that the antibody positivity rate for the N antigen of SARS-CoV-2 increased to 2.1% and 3.9% in those surveys, respectively, indicating the infectious rate of the 5th wave.

## MATERIALS AND METHODS

### Serum samples for seroepidemiologic analysis

The serum samples were collected from individuals who underwent a health check-up at the clinics of Hyogo Prefecture Health Promotion Association, Kobe, Japan. The 1,000 sera collected during the period from July 19 to August 6, 2021 are referred to herein as the August 2021 cohort. The 1,000 sera collected during the period from November 22 to December 8, 2021 are referred to as the December 2021 cohort. No prior information about the individuals from who the sera was collected is described in this study (e.g., vaccination status and SARS-CoV-2 infection history).

### Detection of anti-Spike antibodies by ELISA

Anti-Spike antibodies in the human sera were detected by an anti-S enzyme-linked immunosorbent assay (ELISA) as described in our previous study (manuscript submitted). Briefly, each well of a 96-well ELISA plate (Corning, New York, NY) was coated with 100 ng of Spike protein dissolved in a 100 mM carbonate buffer (pH 9.0) and left at 4°C overnight. After a wash with phosphate-buffered saline (PBS) containing 0.1% Tween 20 (PBST), PBST supplemented with 1% bovine serum albumin was added as a blocking buffer, followed by incubation at 4°C for 2 hr. Each serum serially diluted from 1:40 to 1:5120 in the blocking buffer were added to the plate and then incubated at 37°C for 1 hr.

After a wash with PBST, goat anti-human IgG with conjugated horseradish peroxidase (abcam, Cambridge, MA) diluted 1:10000 with PBST was added, followed by incubation at 37°C for 1 hr. After a wash with PBST, ABTS solution (Roche Diagnostics, Indianapolis, IN) was added, and the plate was incubated at room temperature for 40 min in the dark. The reaction was stopped by adding 100 μl of 1.5% (w/v) oxalic acid dehydrate solution.

The optical density at wavelength 405 nm (OD_405_) was measured using the plate reader Multiskan FC (Thermofisher Scientific, Waltham, MA). We set a cut-off value of 0.3 for the 1:40 dilution to define anti-S positivity according to the values for native sera according to our validation (manuscript submitted). The value of the area under the curve (AUC) was used to evaluate the anti-S antibody titer [21, 22]. The AUCs were calculated for the plot of OD_405_ values according to dilution factors, and an arbitrary value of 1 was given as the width for a twofold dilution step.

### Detection of anti-N antibody by ECLIA

An electrochemiluminescence immunoassay (ECLIA) was conducted using the cobas e801 module (Roche Diagnostics, Rotkreuz, Switzerland) as in our previous study [20]. The Elecsys Anti-SARS-CoV-2 assay kit (Roche Diagnostics) was used for the detection of antibodies against the SARS-CoV-2 nucleocapsid (N). The measurement of anti-N antibody was performed according to the manufacturer’s instructions, and samples with a cut-off index (COI) >1.0 were diagnosed as positive.

### Viruses

The SARS-CoV-2 Biken-2 (B2) strain, which contains the Spike D614G mutation (here called D614G) was provided by BIKEN Innovative Vaccine Research Alliance Laboratories (Osaka, Japan). The whole genome sequence of the B2 strain has been deposited in the DNA Data Bank of Japan (DDBJ) with the accession number LC644163. The SARS-CoV-2 B.1.167.2 Delta variant (GISAID ID: EPI_ISL_2158617) and the B.1.1.529 Omicron variant (GISAID ID: EPI_ISL_7418017) were provided by Japan’s National Institute of Infectious Disease.

### Neutralization assay

The neutralization activity was measured as described [19, 20, 23] using authentic viruses. The use of SARS-CoV-2 viruses was restricted to the BSL3 Laboratory at the Kobe University.

### Statistical analysis

The Kruskal-Wallis test, Dunn’s multiple comparison test, and Mann-Whitney U-test were performed by GraphPad Prism 8 (GraphPad Software, San Diego, CA). In all statistical tests, *P*-values□<0.05 were considered significant.

### Ethics statement

The seroepidemiologic surveillance was approved by the Ethics Committee of Kobe University Graduate School of Medicine (approval code: B2156702). The Hyogo Prefecture Health Promotion Association was also granted approval for study participation under the same Ethics Committee. Information about this retrospective observational study was published on the website of Kobe University Hospital, along with the opportunity to opt out.

## RESULTS

### Large-scale seroepidemiologic surveillance of the anti-N positive rate

The results of our large-scale seroepidemiologic surveillance in Hyogo prefecture, Japan performed in August 2021 and December 2021 are as follows. As many as 1,000 samples in each cohort were collected from individuals who underwent a health check-up at Hyogo Prefecture Health Promotion Association clinics, with no prior information about the individuals’ SARS-CoV-2 infection and vaccination history. The demographic data are summarized in Table 1, and the age distributions are depicted in the left panels of Figures 1A and 1B for the August 2021 and December 2021 cohorts, respectively. These cohorts were composed mainly of persons of 20–69 years old with a smaller number of persons aged 70–80, with few individuals <20 years old and no subjects aged <18 years old.

**Table 1.** Demographic information of the cohorts for the large-scale seroepidemiologic surveillance

|  | August 2021 |  |  | December 2021 |  |  |
| --- | --- | --- | --- | --- | --- | --- |
|  | All | Male | Female | All | Male | Female |
| n (%) | 1,000<br>(100%) | 587<br>(58.7%) | 413<br>(41.3%) | 1000<br>(100%) | 508<br>(50.8%) | 492<br>(49.2%) |
| Age, yrs, median,<br>(range) | 48<br>(19–83) | 51<br>(20–83) | 44<br>(19–78) | 49<br>(18-79) | 42a<br>(19-79) | 49<br>(18-75) |
| Age groups, yrs, n: |  |  |  |  |  |  |
| 18-19* | 4 | 0 | 4 | 3 | 2 | 1 |
| 20–29 | 123 | 41 | 82 | 104 | 48 | 56 |
| 30–39 | 179 | 97 | 82 | 149 | 85 | 64 |
| 40–49 | 243 | 134 | 109 | 257 | 123 | 134 |
| 50–59 | 236 | 153 | 83 | 265 | 123 | 142 |
| 60–69 | 174 | 133 | 41 | 182 | 99 | 83 |
| 70–83† | 41 | 29 | 12 | 40 | 28 | 12 |

\* The August 2021 cohort contains only 19 years old in this age group
† The December 2021 cohort contains up to 79 years old in this age group

**Fig. 1.**
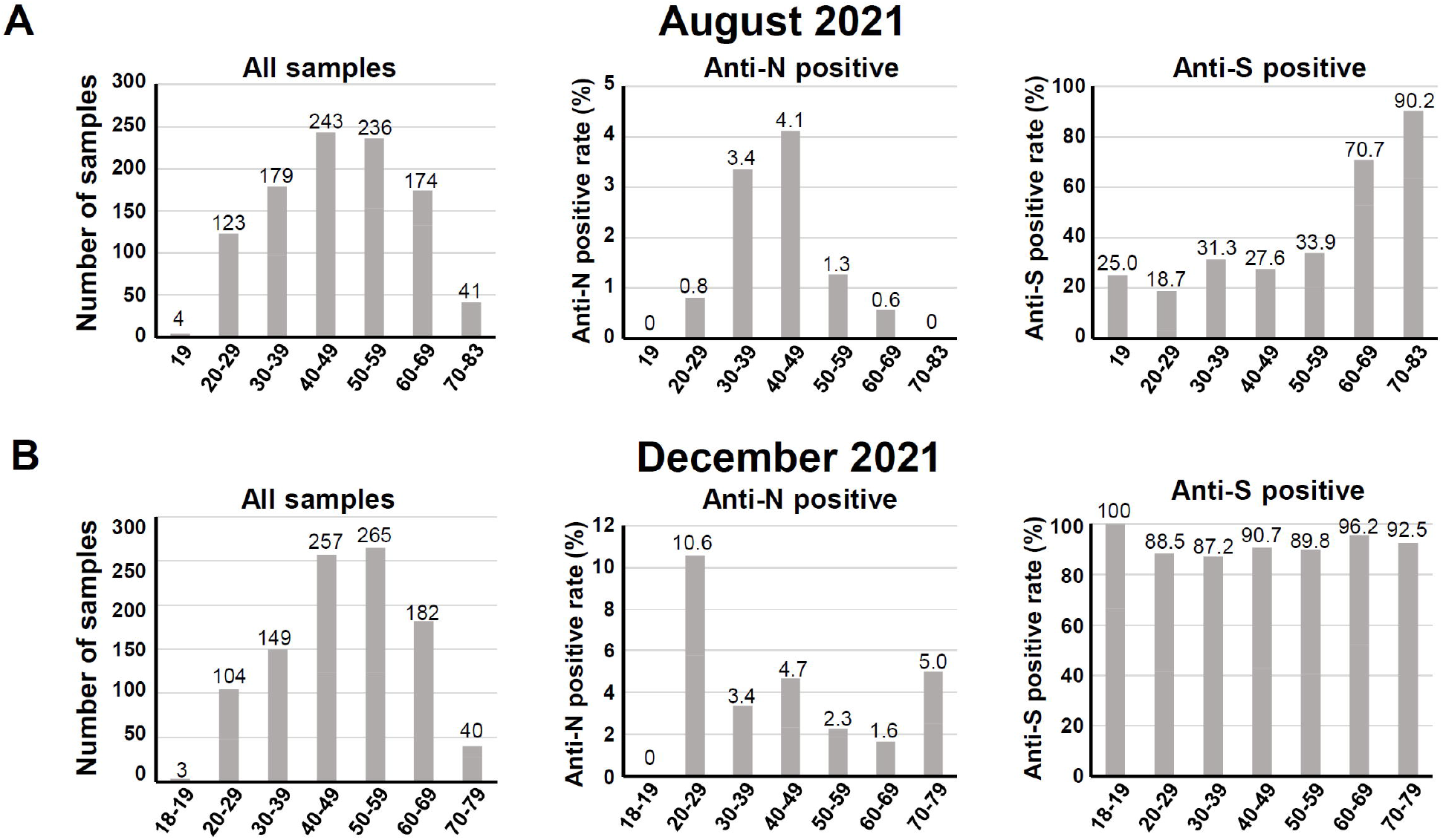
The anti-N and anti-S antibody seroprevalence of the August 2021 and the December 2021 cohorts. **A:** *Left panel:* The age distribution of the August 2021 cohort divided into six age groups: 18–19, 20–29, 30–39, 40–49, 50–59, 60–69, and 70–83 years. *Middle panels:* The anti-N-positive rate and (*right panels*) anti-S-positive rate by the age groups. **B:** The age distribution of the December 2021 cohort: 19, 20–29, 30–39, 40–49, 50–59, 60–69, and 70–79 years.

The SARS-CoV-2 anti-N antibodies which show past infection by SARS-CoV-2 were detected by the ECLIA [20, 24], and 21 of the 1,000 samples (2.1%) in the August 2021 cohort were deemed positive given the cut-off of 1.0, whereas 39 of the 1,000 samples (3.9%) in the December 2021 cohort were positive (Table 2). The age distribution of positive cases is summarized in Table 2 and illustrated in the middle panels of Figure 1A,B for the August and December 2021 cohorts, respectively. In the August 2021 cohort, the anti-N positive rate for the age groups 30–39 and 40–49 were relatively high at 3.4% (6/179) and 4.1% (10/243), respectively, and no positive cases were observed for the age groups 18–19 and 70–83 years (Fig. 1A, middle panel).

**Table 2.** Numbers of anti-N- and anti-S-positive samples by age groups

|  | August 2021 |  |  | December 2021 |  |  |
| --- | --- | --- | --- | --- | --- | --- |
|  | All<br>(n=1,000) | Male<br>(n=587) | Female<br>(n=413) | All<br>(n=1,000) | Male<br>(n=508) | Female<br>(n=492) |
| **SARS-CoV-2 anti-N antibody (ECLIA)** |  |  |  |  |  |  |
| Positive no. by age group, yrs, n |  |  |  |  |  |  |
| 18-19* | 0 | 0 | 0 | 0 | 0 | 0 |
| 20-29 | 1 | 0 | 1 | 11 | 6 | 5 |
| 30-39 | 6 | 5 | 1 | 5 | 5 | 0 |
| 40-49 | 10 | 7 | 3 | 12 | 6 | 6 |
| 50-59 | 3 | 3 | 0 | 6 | 5 | 1 |
| 60-69 | 1 | 1 | 0 | 3 | 1 | 2 |
| 70-83† | 0 | 0 | 0 | 2 | 1 | 1 |
| Positive no. in all subjects, n (%) | 21<br>(2.1%) | 16 (2.7%) | 5<br>(1.2%) | 39<br>(3.9%) | 24<br>(4.7%) | 15<br>(3.0%) |
| **SARS-CoV-2 anti-S antibody (ELISA)** |  |  |  |  |  |  |
| Positive no. by age group, yrs, n |  |  |  |  |  |  |
| 18-19* | 1 | 0 | 1 | 3 | 2 | 1 |
| 20-29 | 23 | 4 | 19 | 92 | 43 | 49 |
| 30-39 | 56 | 30 | 26 | 130 | 74 | 56 |
| 40-49 | 67 | 28 | 39 | 233 | 110 | 123 |
| 50-59 | 80 | 48 | 32 | 238 | 107 | 131 |
| 60-69 | 123 | 92 | 31 | 175 | 95 | 78 |
| 70-83† | 37 | 26 | 11 | 37 | 26 | 11 |
| Positive no. in all subjects, n (%) | 387<br>(38.7%) | 228<br>(38.8%) | 159<br>(38.5%) | 908<br>(90.8%) | 457<br>(90.0%) | 449<br>(91.3%) |

\* The August 2021 cohort contains only 19 years old in this age group
† The December 2021 cohort contains up to 79 years old in this age group

Notably, the December 2021 cohort exhibited the noticeable positive rate 10.6% (11/104) for the age group 20–29 (Fig. 1B, middle panel). In contrast to the August 2021 cohort, the oldest age group (i.e., 70–79 yrs) of the December 2021 cohort showed the positive rate 5.0% (2/40). The positive rate for age groups 30–39 and 40–49 were not largely changed at 3.4% (5/149) and 4.7% (12/257), respectively. These results demonstrate that the infection of the younger (20–29 yrs) and older (70–79 yrs) groups increased during the pandemic’s 5th wave.

### Large-scale seroepidemiologic surveillance of the anti-S positive rate

The anti-S antibodies of the August and December cohorts were analyzed by the anti-S ELISA, which included the vaccination and/or infection history. With the cut-off of 0.3 for the 40-fold serum dilution, the positivity rates were 38.7% (387/1,000 sera) and 90.8% (908/1,000 sera) in the August and the December cohorts, respectively (Table 2). The age distribution of anti-S-positive cases is summarized in Table 2 and shown in the right panels of Figures 1A and B. In the August cohort, the anti-S positive rates for the age groups 60–69 and 70–83 were relatively high at 70.7% (123/174) and 90.2% (37/41), respectively (Fig. 1A, right panel). The December cohort demonstrated a flattened positive rate at a high level (87.2%–100%) across all of the age groups tested (Fig. 1B, right panel).

As expected, all of the anti-N-positive sera were also anti-S-positive, with one exception for the August cohort and another for the December cohort.

### Quantitative analysis of the anti-S antibody in the two cohorts

We quantitatively analyzed the anti-S positive sera in the August 2021 and December 2021 cohorts to estimate and compare the cohorts’ anti-S antibody levels. The quantitative distributions of the anti-S antibody titer are shown in Figure 2A. The August 2021 cohort’s distribution showed a significantly higher level of AUCs with the median 13.8 compared to that of the December 2021 cohort with the median 8.8 (Mann-Whitney U-test).

**Fig. 2.**
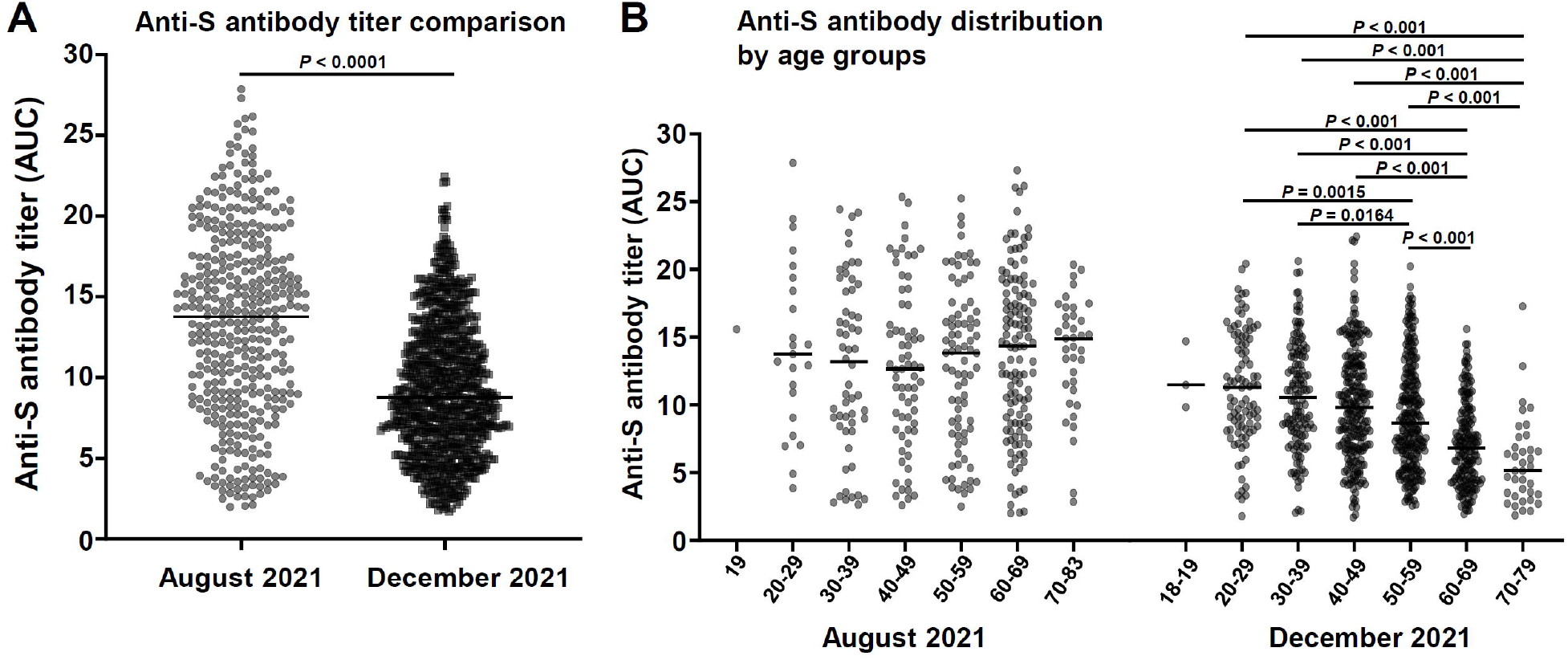
Quantitative evaluation of the anti-S antibody amount by the anti-S ELISA. **A:** The distributions of the anti-S ELISA AUC values were compared between the August 2021 and December 2021 cohorts. *P-*value, Mann-Whitney U-test. *Black bars:* medians. **B:** The distributions in panel A plotted separately for the age groups. The Kruskal-Wallis test was performed for the multiple comparison among age groups in the August and December cohorts. The *P-*values were obtained by the subsequent Dunn’s multiple comparison test for significant data (<0.05). *Black bars:* medians.

We further analyzed the distributions of the anti-S antibody titer by separating the age groups (Fig. 2B). There was no significant difference in the distribution among age groups in the August cohort by the Kruskal-Wallis test, whereas the distribution in the December cohort showed significant differences among the age groups, showing a clear tendency of decreased anti-S antibody levels in the older age group (70–79), indicating that the lower anti-S antibody titer did affect infection.

### Neutralization activity of the anti-S-positive sera from the August 2021 cohort

The neutralization antibody titers of the anti-S-positive subset (n=387) from the August 2021 cohort were quantitatively evaluated. Figure 3A depicts the distributions of neutralization antibody titers against D614G and the Delta variant. Although the distributions appeared similar with the same median value, i.e., 8, the Mann-Whitney U-test result indicated that the neutralizing titers for D614G were significantly higher than those for the Delta variant. The neutralization positivity rate in the anti-S-positive subset also showed a similar result in that there were slightly higher rates for D614G and Delta at 85.5% and 77.3%, respectively (Fig. 3B), indicating the efficacy of the two-dose vaccine for the Delta variant as well.

**Fig. 3.**
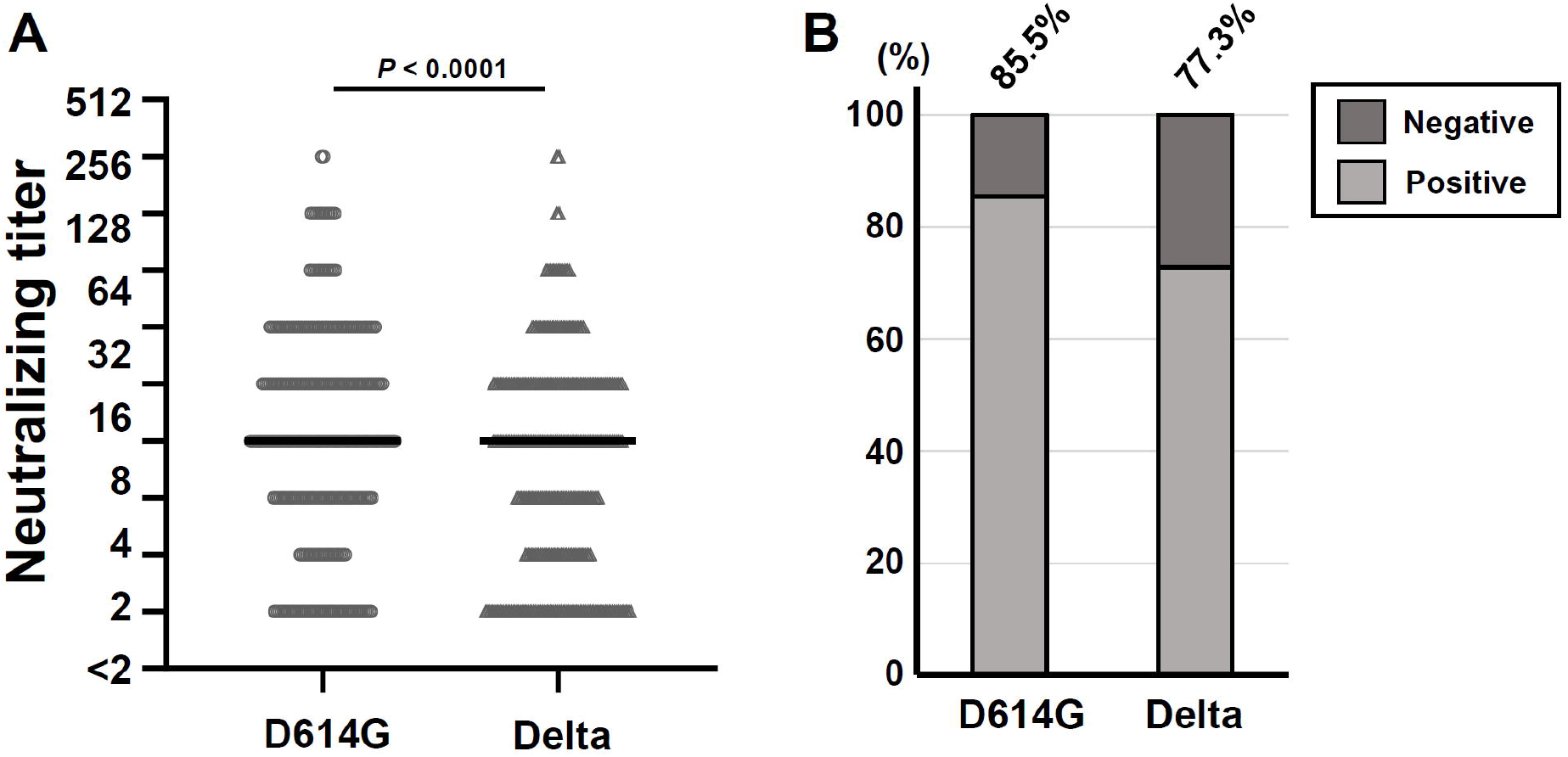
**A:** Neutralization antibody titers of the anti-S-positive subset of the August 2021 cohort plotted for D614G and the Delta variant. *Black bars:* medians. *P-*value, Mann-Whitney U-test. **B:** The neutralization-positive rates for D614G and the Delta variant were evaluated from the data shown in panel A, with the titers <2 deemed negative.

### Neutralizing activities of the sera collected in December 2021 against the Delta and Omicron variants

We next analyzed the neutralization positivity rate against the Delta and Omicron variants in all 1,000 sera of the December 2021 cohort. The neutralization positivity rate against the Delta variant was 78.7% (787/1,000) in the December 2021 cohort, indicating effective social immunity (Fig. 4A, Table 3). In contrast, the neutralization positivity rate against the Omicron variant was quite low at 36.6% (366/1,000) (Fig. 4A, Table 3), implying the vulnerability of the cohort against infection by the Omicron variant.

**Table 3.** Neutralization positive rate of the sera collected on December 2021

|  | Neutralization positive number, (%) |  |
| --- | --- | --- |
|  | Delta | Omicron |
| by age groups, yrs, |  |  |
| 18-19 (n=3) | 3 (100%) | 3 (100%) |
| 20-29 (n=104) | 86 (82.7%) | 59 (56.7%) |
| 30-39 (n=149) | 120 (80.5%) | 62 (41.6%) |
| 40-49 (n=257) | 216 (84.0%) | 104 (40.5%) |
| 50-59 (n=265) | 209 (78.9%) | 91 (34.3%) |
| 60-69 (n=182) | 132 (72.5%) | 41 (22.5%) |
| 70-79 (n=40) | 21 (52.5%) | 6 (15.0%) |
| in all subjects (n=1,000) | 787 (78.7%) | 366 (36.6%) |

**Fig. 4.**
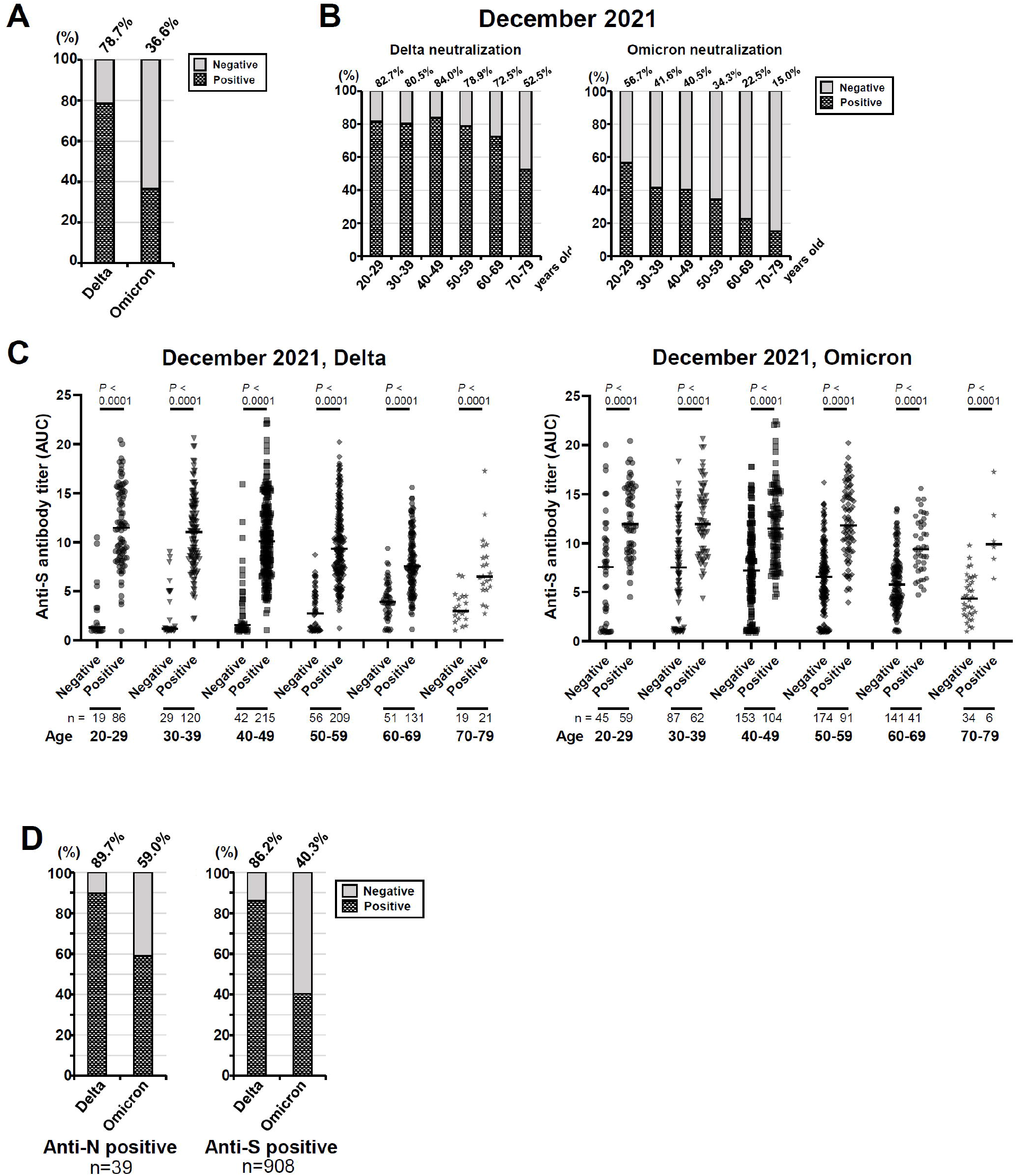
Neutralization activity of the sera collected in December 2021 against the Delta and Omicron variants. **A:** The neutralization-positive rates of the entire December 2021 cohort (n=1,000) plotted for the Delta and Omicron variants. **B:** The neutralization-positive rates for the Delta and Omicron variants by age groups. The data in panel A were reanalyzed as in Fig. 2B. **C:** Comparison of the anti-S antibody amount between the neutralization-positive and -negative groups. The complete set of 1,000 sera of the December 2021 cohort were subgrouped according to result of the neutralization assay against the Delta variant and the Omicron variant, and the anti-S antibody titers were plotted. *P*-value, Mann-Whitney U-test. *Black bars*: medians. **D:** The neutralization-positive rate calculated for the anti-N-positive (n=39) and anti-S (n=908) subsets obtained by extracting the data from those shown in panel A.

A further analysis of the same data with a separation of age groups revealed that the neutralization positivity rate against the Delta variant tended to be lower in older age groups (Fig. 4B, Table 3). The same trend was also observed in the neutralization positivity rate against the Omicron variant (Fig. 4B, Table 3), and in the oldest age group (70–79 yrs) only 15.0% of the sera demonstrated the neutralization activity.

The comparison of the anti-S antibody titers between the neutralization-positive and -negative sera revealed that the neutralization-positive group had clearly higher titers of anti-S antibody in both the Delta and Omicron cases (Fig. 4C), supporting our speculation that the neutralization activity of the sera was attributed to the anti-S antibodies.

Lastly, we focused on the anti-N-positive (n=39) and anti-S positive (n=908) subsets of the December 2021 cohort. The neutralization-positive rate against the Delta and the Omicron variants for the anti-N positive subset were 89.7% and 59.0%, respectively (Fig. 4D). These values were slightly higher than those for the entire set of 1,000 sera or those for the anti-N negative subset (78.3% and 35.7% for the Delta and the Omicron variants, respectively), indicating superior immune responses in the individuals with a history of SARS-CoV-2 infection. The neutralization positive rate for the anti-S positive subset were 86.2% and 40.3% against the Delta and the Omicron variants, respectively.

## DISCUSSION

In combination with other methods used to understanding the spread of infectious diseases such as the polymerase chain reaction (PCR)-based diagnosis and antigen tests, periodic cross-sectional seroepidemiologic surveillance is a powerful approach. Our present surveillance in Japan’s Hyogo prefecture revealed that the anti-N-positive rate, which represents the SARS-CoV-2 infection rate, was 2.1% in the August 2021 cohort and 3.9% in the December 2021 cohort. In our October 2020 serosurveillance [20], the anti-N-positive rate by the same ECLIA method for 1,000 sera from the same Hyogo Prefecture Health Promotion Association clinics was 0.4%. The increase in the anti-N-positive rate in Hyogo prefecture was well synchronized with the cumulative number of the COVID-19 cases based on PCR results (Fig. 5). The increased rate of anti-N seroprevalence was 0.17% per month from October 2020 to August 2021 and 0.45% per month from August 2021 to December 2021. The steep increase during the latter period is accounted for by the spread of the Delta variant in Japan’s so-called ‘5th wave’ (Suppl. Fig. S1).

**Fig. 5.**
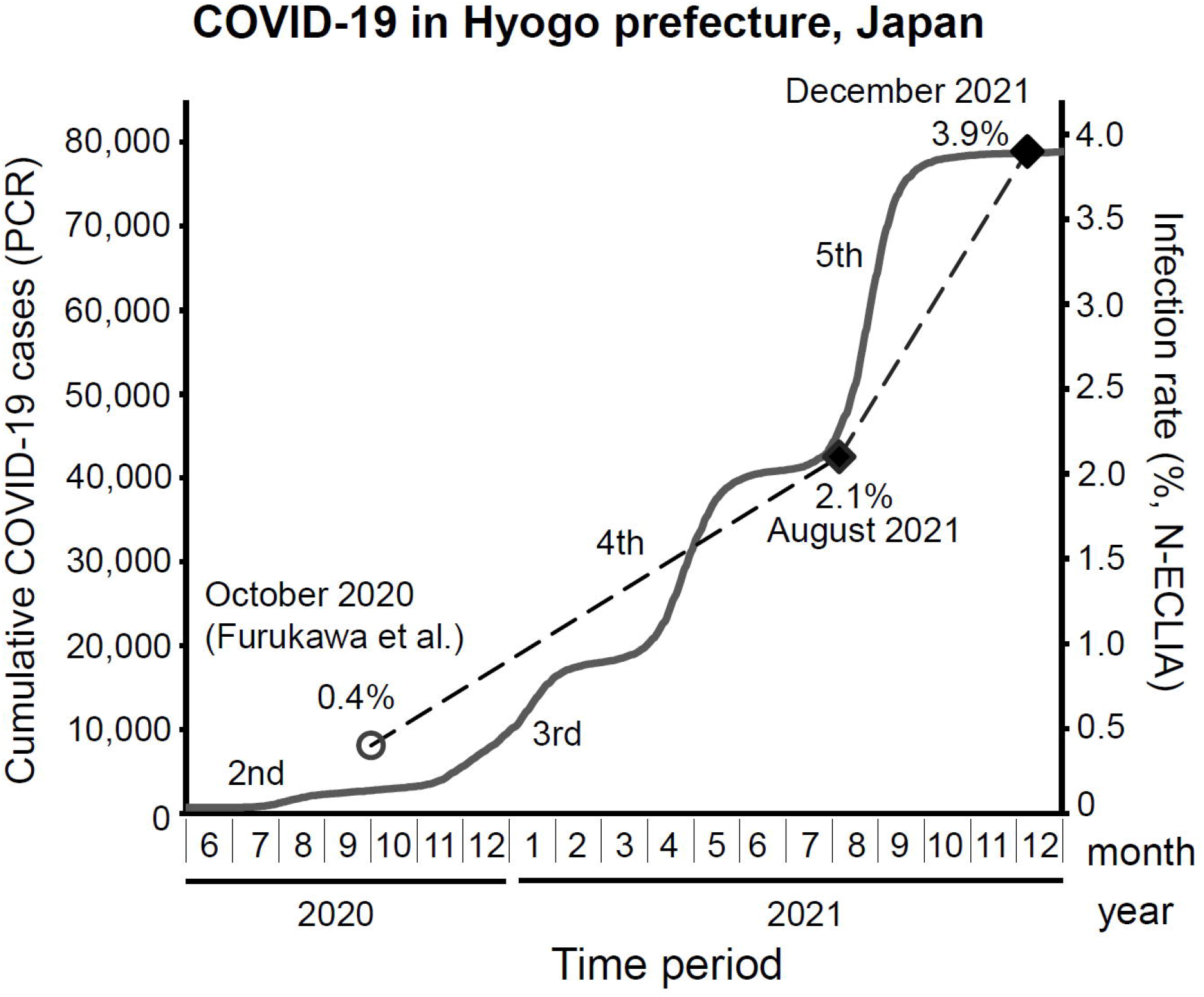
The COVID-19 situation in Hyogo prefecture, Japan. The cumulative infection cases reported based on the PCR diagnosis [1] are plotted as the *solid line*, and the infection rates obtained by our serosurveillance surveys are plotted as the *dashed line*. The anti-N-positive rate shown by the ECLIA for the October 2020 cohort was obtained from our previous study [20]. The rapid increase in the number of COVID-19 cases, i.e., the so-called 2nd to 5th waves in Japan, are also indicated.

We suspected that the SARS-CoV-2 infection rate estimated by the seroprevalence of anti-N antibody would be higher than the rate indicated by the PCR-based diagnoses due to the existence of asymptomatically or mildly infected individuals whose cases may not have been detected since they never underwent testing by PCR analysis or antigen test. Indeed, the anti-N ECLIA-based infection rates revealed in the present analyses, i.e., 2.1% and 3.9% for August and December 2021, were respectively higher than the rates 0.85% (45,789 cases/5.4 million people in Hyogo prefecture as of August 6, 2021) and 1.5% (78,718 cases/5.4 million people in Hyogo prefecture as of December 8, 2021) estimated from the reported cases based on PCR diagnoses [1]. The fold difference was nearly constant for both periods, reflecting the synchronized increases in the anti-N-positive rate and the reported cases (Fig. 5), i.e., 2.5-to 2.6-fold lower for the PCR-based estimation.

Even if we compensate for the biased age distribution in our cohorts (which were composed mainly of 20-to 80-year-olds), it is likely that the PCR-based infection rates are approx. 2-fold lower than the infection rate revealed by our serosurveillance, given the matched population 4.1 million for 20- to 80-year-olds and the finding that no less than 80% of COVID-19 cases in Japan are among 20- to 80-year-olds [25]. This is consistent with the report that a substantial portion of SARS-CoV-2-infected individuals are asymptomatic [26], leading to the inaccuracy of surveillance rates determined solely by PCR diagnosis.

Two-dose COVID-19 vaccination has been intensively promoted in Japan from May to December 2021, and approx. 74% population has finished the two-dose vaccination (as of January 6, 2022; [12]). This is a relatively high rate considering the global average (46%) in January 2022 (January 6, 2022; [27]). Our anti-S seroprevalence result for the August 2021 cohort (38.7%) represents the build-up period of social immunity (Table 2, Fig. 1A), and it is well consistent with Hyogo prefecture’s single and two-dose vaccination rates at 32.79% and 42.05%, respectively as of August 6, 2021. In December 2021, when the vaccination rate had reached a plateau in Japan, our survey revealed an extremely high seropositive rate, i.e., 90.8% (Table 2, Fig. 1B). This result indicates that the vaccinations in Japan (*i*) have established herd immunity, and (*ii*) may have contributed to the current COVID-19 situation having calmed down in December 2021 (Suppl. Fig. S1, Fig. 5).

The anti-S seroprevalence observed in the present study is clearly higher compared to the reported single- and two-dose vaccination rates in Hyogo prefecture as of December 8, 2021 at 73.09% and 74.01%, respectively. This difference might be accounted for by the bias in age groups of the present cohorts which contained very few people <20 years old, whose vaccination rate has been relatively low compared to those of other age groups [28]. In addition, our cohorts consisted of individuals who underwent a health check-up, and this could have contributed to a biased selection of socially active and health-conscious people.

Our comparison of the anti-N and anti-S seropositive rates among the age groups revealed differing tendencies between the August 2021 and December 2021 cohorts (Fig. 1A,B). The August 2021 cohort□—□which was enrolled during a period of vaccination progression and the beginning of the Delta variant’s rampage□—□represents competing effects of the vaccinations and the Delta variant. The high anti-S-positive rate for the age groups 60–69 and 70–83 years observed herein is explained by the priority vaccination for elderly individuals (>65 yrs) in Japan (Fig. 1A). Interestingly, the high anti-S-positive rate might be related to the relatively low infection rate (low anti-N-positive rate) in the present study’s elderly age groups (Fig. 1A), indicating the vaccines’ efficacy.

In contrast, the December 2021 cohort was enrolled during the period in which the ‘fully vaccinated’ rate had reached a plateau and the increase in the number of COVID-19 cases due to the Delta variant had dropped to a low level (Suppl. Fig. S1). The anti-N-positive rate was increased in the 20–29 and 70–79 age groups in this cohort (Fig. 1B). Although the anti-S-positive rate of the December 2021 cohort showed no apparent difference among age groups (Fig. 1B), our quantitative analysis of the anti-S antibody titers suggested that the increased infection for the 70- to 79-year-olds was the result of the decreased anti-S antibody titers for the elderly age groups (Fig. 2B). Indeed, the anti-S antibody level was significantly lower for the December 2021 cohort compared to that for the August 2021 cohort (Fig. 2A), reflecting the decline of acquired immunity after vaccination. Regarding the increase in the positive rate of anti-N antibody in the 20–29 age group, we speculate that this increase might reflect increased social activity among these younger individuals.

As shown in Figure 4A, the neutralization assay using the SARS-CoV-2 viruses revealed that the December 2021 cohort had a high neutralization-positive rate for the Delta variant, i.e., 78.7%. This result demonstrated the two-dose vaccination suppressed the spread of the Delta variant, leading to a transient convergence of the COVID-19, resulting in the relatively low infection rate of 3.9%.

The emergence of the Omicron variant is a key event worldwide, as is the subsequent spread of the Delta variant. As of December 2021, European countries and U.S. have been invaded by the Omicron variant, which has also been detected across Japan. Our neutralization assay against the Omicron variant for the December 2021 cohort, which represents the current Japanese vaccinated population, demonstrated a quite low positive rate, 36.6% (Fig. 4A) which appears to be due at least in part to several mutations that occurred in the S protein [29-31]. We thus contend that the three-dose vaccination to boost the immunity against Omicron is needed to prevent and suppress the further spread of the Omicron variant. Our present findings also demonstrate the efficacy of vaccines for SARS-CoV-2 suppression. The booster should be required, in order to increase the population’s immunity to the novel variant Omicron.

## Data Availability

There is no available data referred to the manuscript.

## Acknowledgments

We thank Kazuro Sugimura MD, PhD (Superintendent, Hyogo Prefectural Hospital Agency and Professor, Kobe University) for his full support of this study. We express our sincere gratitude for the cooperation and participation of the staff of the Hyogo Prefecture Health Promotion Association. We also thank researchers of the Division of Respiratory Medicine, Department of Internal Medicine, Kobe University Graduate School of Medicine. We thank BIKEN Innovative Vaccine Research Alliance Laboratories for providing the SARS-CoV-2 B2 strain used herein as the D614G variant. We also thank the National Institute of Infectious Disease Japan for providing the SARS-CoV-2 B.1.167.2 variant and the B.1.1.529 variant used as the Delta variant and the Omicron variant, respectively. YK was supported by the Taniguchi Memorial Scholarship program provided by the BIKEN Foundation. SS and LHT were supported by Japanese Government (Monbukagakusho: MEXT) Scholarships.

## Contributions

ZR contributed to all of the experiments and analyses of the data. ZR, LHT, KF, YK, and SS performed the neutralization assay. MN, KF, JA and YM supported the maintenance of the Biosafety Level 3 laboratory. ZR, LHT, KA, and NH performed the S ELISA measurement. KU, KM, IS, and JS performed the N ECLIA measurement. NG, KT, and MY performed the large-scale protein expression and purification of the Spike antigen for the S ELISA measurement. TN collected the sera and managed their attributed data. YM conducted and supervised the study. ZR, LHT, FK, MN, JA, and YM summarized the entire data set. The draft of the manuscript was written by ZR, LHT, MN, and YM. All authors approved the final version of the manuscript.

