## Supplementary Figure S1 for "Large-scale cross-sectional seroepidemiologic study of COVID-19 in Japan: Acquisition of herd immunity and the vaccines’ efficacy"

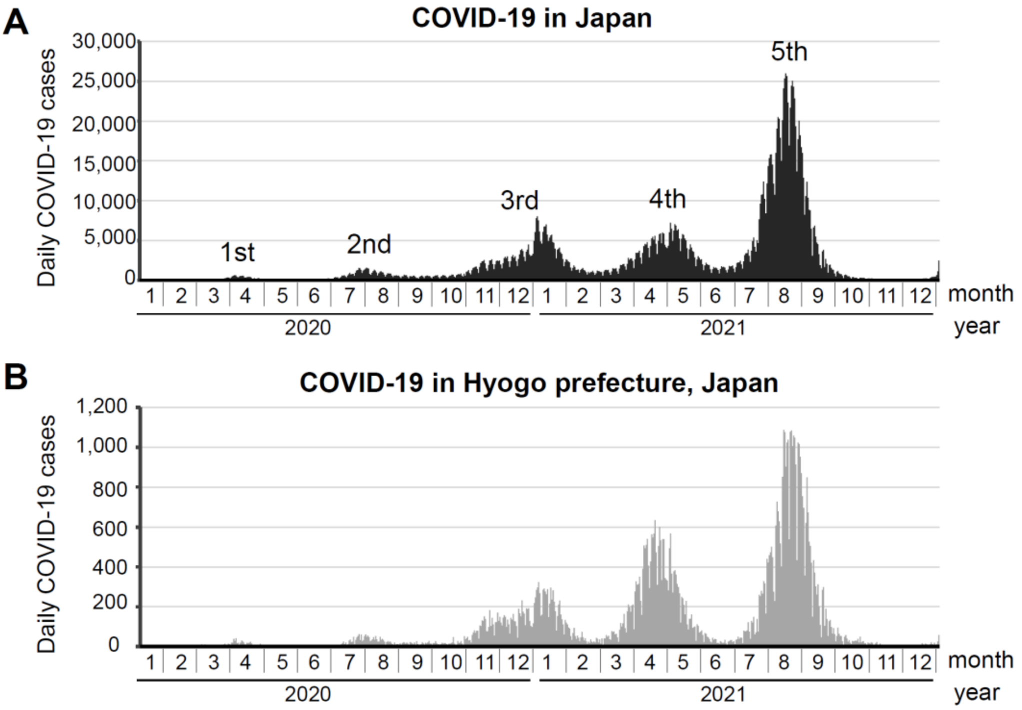


Data were obtained from https://www.mhlw.go.jp/stf/covid-19/open-data_english.html and modified for presentation

**Suppl. Fig. S1.** The COVID-19 situation in Japan (nationwide) and in Hyogo prefecture. The daily COVID-19 cases reported based on PCR diagnoses in Japan **(A)** and in Hyogo prefecture **(B)** are plotted from January 2020 to January 2022. The data were obtained from the website provided by Japan's Ministry of Health, Labour and Welfare (https://www.mhlw.go.jp/stf/covid-19/open-data_english.html) [1] and modified. The surges of SARS-CoV-2 spread that have occurred in Japan, called the 1st to 5th waves, are indicated in panel A.
